# Six recommendations for improving care and support in advanced COPD: evidenced from a population-based, mixed method, multiple-perspective, longitudinal study

**DOI:** 10.64898/2026.09.29.26364287

**Authors:** Morag Farquhar, Gail Ewing, Caroline Moore, A. Carole Gardener, Hanne Holt Butcher, Peter Burge, Tom Ling, Living with Breathlessness Study Team

## Abstract

**Background:** Chronic obstructive pulmonary disease (COPD) is a progressive condition with high symptom burden. Advanced COPD management should relieve symptoms, optimise daily functioning and reduce carer burden, yet there is limited enactment of policy-advocated person-centred approaches to care and support. The Living with Breathlessness Study sought to establish evidence-based recommendations to inform a new paradigm to improve care and support in advanced COPD.

**Methods:** Comprehensive multiple-perspective longitudinal evidence was collected on care, support, functioning and needs in advanced COPD from a population-based cohort of over 500 patients, their unpaid/family carers and health care professionals through semi-structured interviews and validated outcome measures over an 18-month period. Quantitative data analysed using standard descriptive statistics, numerical and univariate analyses, and qualitative data using a Framework approach.

Recommendations were then generated through three linked strategies: (1) an expert multidisciplinary group, (2) a stakeholder workshop and (3) a stakeholder survey.

**Results:** Six inter-related recommendations emerged, linked by the concept of proactive person-centred care, supported by action points to enable delivery. Recommendations in brief: (1) stop focusing on the challenge of prognostication as a barrier to meeting need, (2) change targets to incentivise person-centred care, (3) enable identification of patient support needs, (4) identify and support carers, (5) identify and respond to psychological morbidity, (6) change societal understandings of COPD, breathlessness, palliative care and informal carers. The recommendations garnered significant support from stakeholders, with caution regarding ease of implementation.

**Conclusions:** Enactment of the resulting six evidence-based recommendations, through their action points, could inform a new paradigm to improve care and support in advanced COPD recognising the slow relentless progression of non-malignant disease, and its effect on patients and their carers. They hold potential for other patients and unpaid/family carers who are living in between conventional disease management and end-of-life care, and for those HCPs striving to support them.

## INTRODUCTION

Chronic obstructive pulmonary disease (COPD) is a common chronic progressive respiratory long-term condition with high symptom [1,2] and unpaid/family carer burden [3,4]. There are over 250 million cases of COPD worldwide and it is the third leading cause of death globally, with the burden from the disease predicted to continue to rise [5,6]. When COPD is advanced, patients can become housebound with severe breathlessness [7–11] suffering slow decline in quality of life, punctuated by acute exacerbations with only partial recovery until final crisis [12].

Advanced COPD management should relieve symptoms, optimise daily functioning and reduce carer burden [13,14]. However, there is limited enactment of policy-advocated person-centred approaches to care and support [15–19]. Failure to address care and support needs of this group is known [15,20,21] including a lack of information about COPD, practical assistance, psychological support, social engagement and end-of-life care. National strategy documents state that quality end-of-life care should be available to patients with any condition [22,23], however prognostication is difficult and access to palliative care remains poor for COPD compared to cancer [7,24,25]. Further, the Ambitions for Palliative and End-of-life Care national framework [26] includes long-term conditions in its priorities, yet in advanced non-malignant disease we continue to rely on pathways and service frameworks developed for cancer with its different, more predictable, trajectory [27].

Limited knowledge of support needs and preferences for care in advanced COPD has created uncertainty in the timing and type of relevant effective supportive interventions. An alternative approach to the cancer framework is required, based on recognition of the slow relentless progression of non-malignant disease, its effect on patients, and on those unpaid/family carers who support them. The Living with Breathlessness Study programme (LwB) sought to establish evidence-based recommendations to inform a new paradigm to improve care and support in advanced COPD. This paper reports those evidence-based recommendations. To our knowledge, at the time of its conduct, LwB was the largest UK population-based, mixed method, multiple-perspective, longitudinal study of care and support in advanced COPD.

## METHODS

We collected mixed method longitudinal evidence on experiences, preferences and outcomes of care and support from the multiple-perspectives of patients with advanced COPD, their unpaid/family carers (family and friends who support them), and patient-nominated key health care professionals (HCPs), between 2013 and 2016, through the multi-component LwB programme. A population-based cohort of over 500 patients with advanced COPD was recruited to the programme through 123 primary care practices in the East of England and South London against clinician-defined inclusion criteria for advanced COPD and study-appropriate exclusion criteria (see Box 1). Recruiting practices were targeted to maximise variability.

#### Box 1: Living with Breathlessness Study programme patient inclusion and exclusion criteria

|  |
| --- |
| **<u>Box 1: Living with Breathlessness Study programme patient inclusion and exclusion criteria</u>** |
| **Inclusion criteria: COPD patients registered with primary care practices with *any two* of:** |
| • FEV1<30%<br>• 2+ exacerbations requiring prednisolone or antibiotics in previous year<br>• admission for COPD in previous 2 years<br>• long term oxygen therapy<br>• cor pulmonale<br>• MRC Dyspnoea scale 4+ |
| **Exclusion criteria (any):** |
| • serious mental health problem<br>• serious learning difficulty<br>• active cancer<br>• active alcoholism |
| **Additional note provided to recruiting practices:** Patients in this study are expected to be older individuals, have serious lung disease, and possibly be housebound, so GPs are asked not to exclude patients for these reasons. |

Evidence was collected through three linked component studies outlined in Box 2.

#### Box 2: Living with Breathlessness Study programme three linked evidence-generating component studies

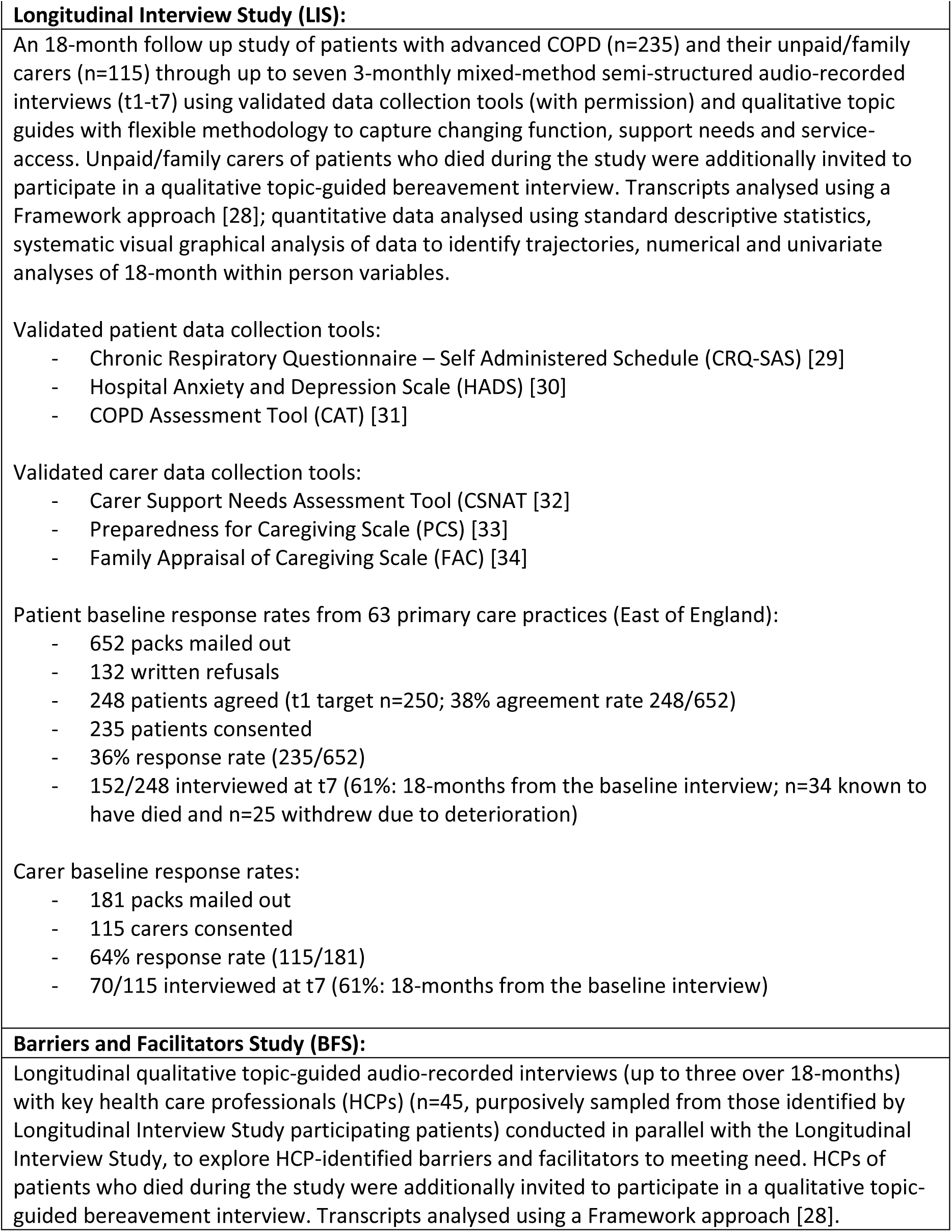
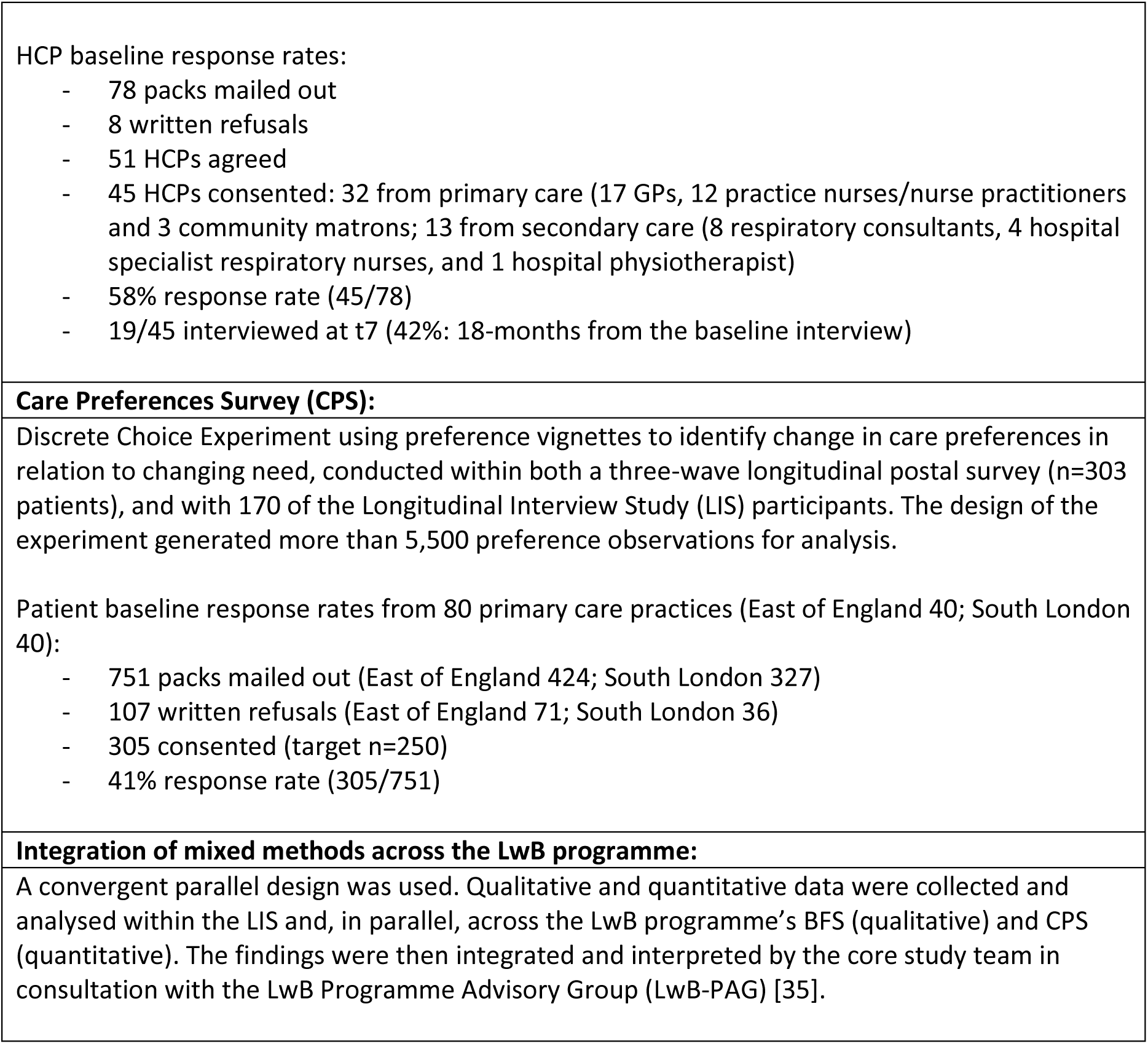

To enable consideration of representativeness, the key baseline characteristics of participating LIS patients and carers are summarised in Box 3.

#### Box 3: Key baseline characteristics of participating LIS patients and carers

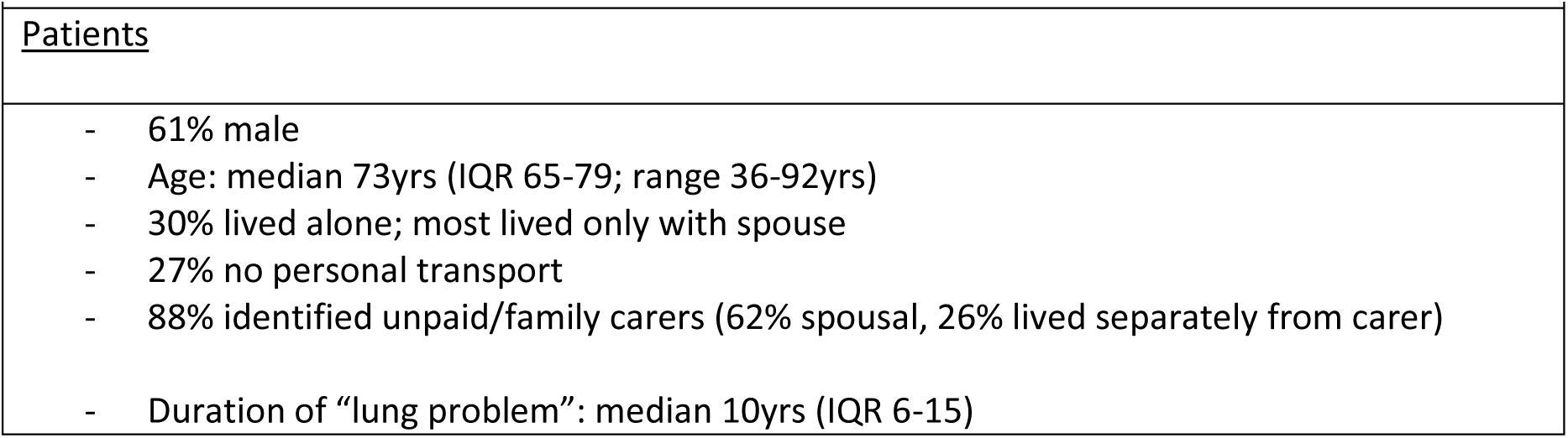
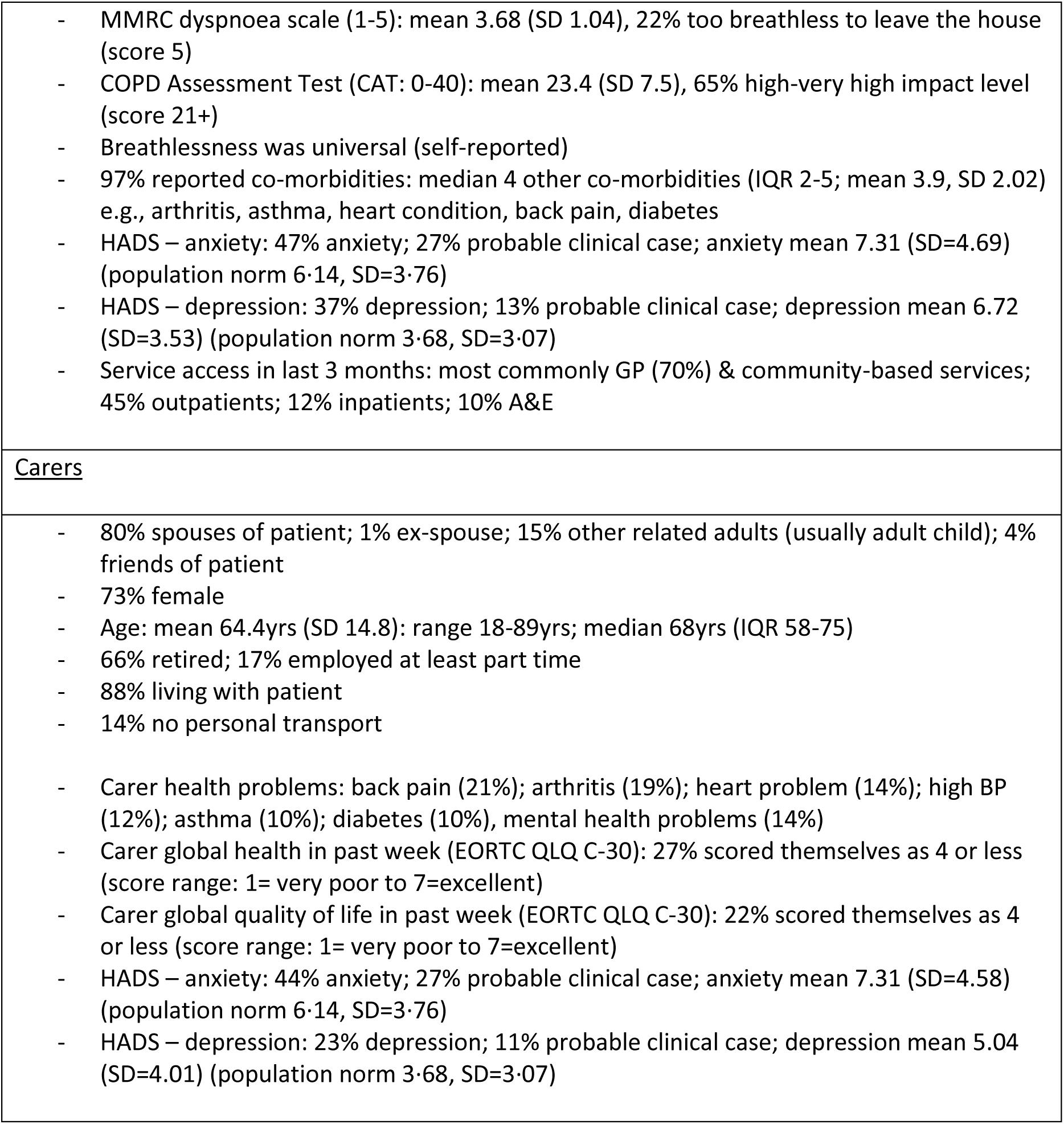

The recommendations resulting from the study were generated through three linked strategies: (1) an expert multidisciplinary group, (2) a stakeholder workshop and (3) a stakeholder survey. As the expert multidisciplinary group, the Living with Breathlessness Study Programme Advisory Group (LwB-PAG) iteratively reviewed emergent study findings at a series of LwB-PAG meetings to inform the development of draft recommendations. The LwB-PAG included clinical experts in COPD (across primary and secondary care, medicine and nursing) palliative care, and carer support as well as methodological experts (n=14), plus LwB’s core study team members (Farquhar, Gardener, Moore,

Holt Butcher). The 15 emergent recommendations were then debated at a final LwB-PAG, resulting in their reformulation as six overarching recommendations with associated action points. A stakeholder workshop was then held at the Royal College of General Practitioners (London, 27th October 2015) to debate the suitability, feasibility, and acceptability of each of the six recommendations. Workshop participants (n=15) were recruited from relevant professional organisations, Royal Colleges, and patient organisations. An online survey, with distribution facilitated by relevant professional organisations, then gathered national stakeholder views (n=83) on whether the recommendations would improve care for people with advanced COPD and the ease of their implementation in their place of work [36].

Ethical approval was obtained from the National Research Ethics Service (NRES) Committee East of England - Cambridge South (Reference number 12/EE/0163). All participants provided written informed consent.

### Patient and Public Involvement

Patients with advanced COPD and their carers were consulted extensively in the development of the LwB programme through meetings with the British Lung Foundation’s two local Breathe Easy groups for Cambridge and rural Fenland, plus individual patients and carers at both individual and group meetings and via email (user preference). At each meeting the LwB programme was presented, discussed and consensus achieved on the best ways take its development forward. This informed the acceptability and feasibility of the programme’s design and its recruitment strategy. Specifically, these consultations advised on LwB programme recruitment materials (e.g., identifying the need to remove the word ‘advanced’ from programme literature for patients and carers) and anti-attrition strategies, and reaffirmed the need for skilled, trained and supported research staff. Patient groups were involved in reviewing and piloting data collection instruments (particularly for the Discrete Choice Experiment) and methods.

## RESULTS

Six evidence-based recommendations for improving care and support in advanced COPD emerged, together with action points to enable their delivery (Table 1). The six recommendations target different stakeholders and operate at person, system, and societal levels. The recommendations are inter-related, working together to enable delivery of proactive, supportive, person-centred care in advanced COPD (Fig. 1).

**Fig 1:**
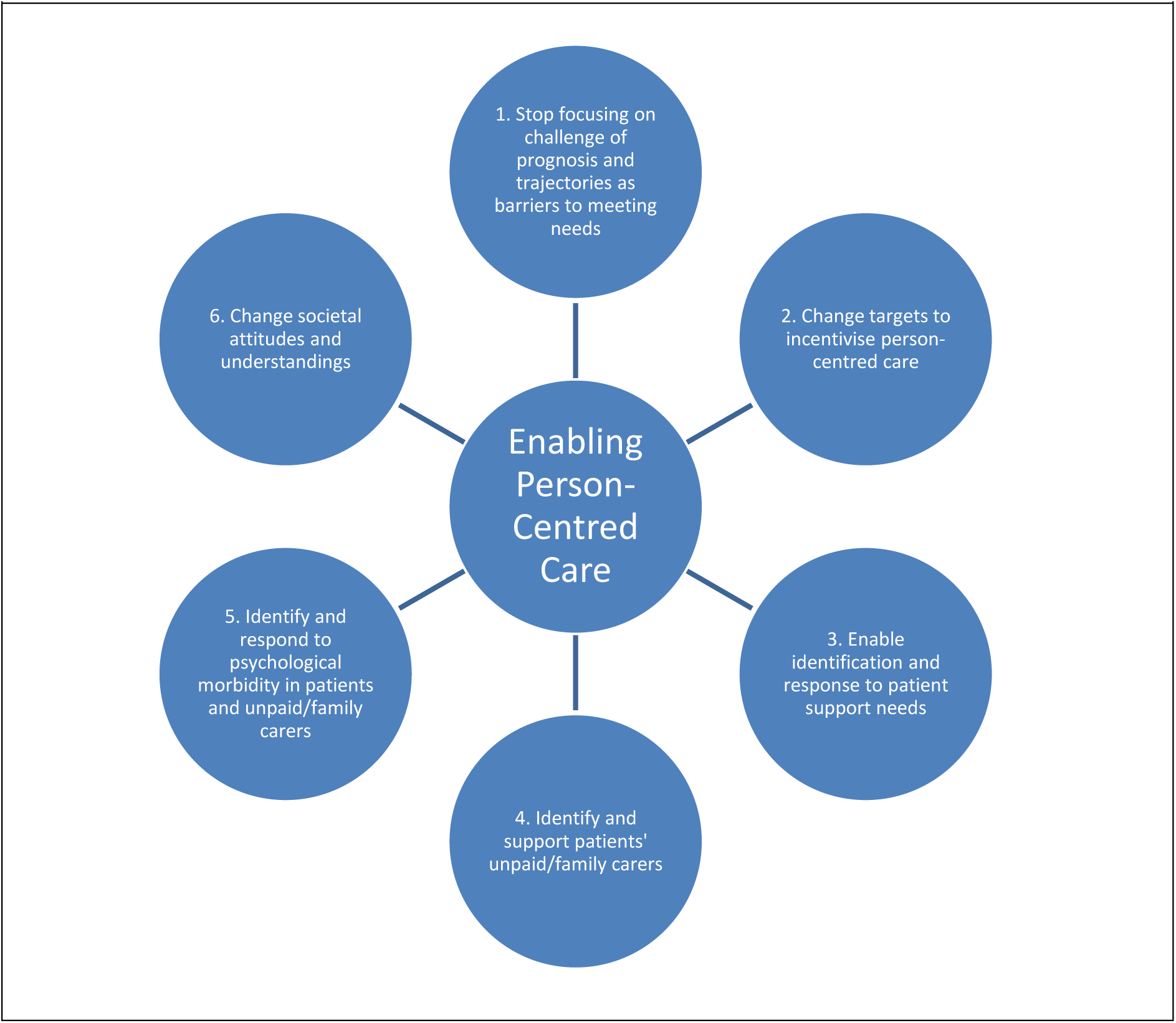
Six inter-related evidence-based recommendations for improving care and support in advanced COPD

**Table 1:** Six evidence-based recommendations, and associated action points, for improving care and support in advanced COPD.

|  |  |
| --- | --- |
| **Six evidence-based recommendations** | **Actions points** |
| 1. Stop focusing on the challenge of prognosis and unpredictability of trajectories as barriers to meeting needs | 1.1 – Enable generalist health care professionals (HCPs) to think differently about access to appropriate care and support, changing the drivers for provision and referral from prognosis and trajectories to patient and unpaid/family carer need<br>1.2 – Support generalist HCPs to make timely referrals to specialist palliative care when required, driven by patient and unpaid/family carer need<br>1.3 – Enable generalist HCPs to provide appropriate non-specialist palliative care to patients and their unpaid/family carers – specialist palliative care is important, but palliative care is not just the remit of specialists – this could be achieved through HCP training across the generalist-specialist interface<br>1.4 – Address the tendency of policy documents and directives to equate palliative care with specialist palliative care, and to equate palliative care with end-of-life care – both are unhelpful<br>1.5 – Change the language we use in relation to future care planning |
| 2. Change targets to incentivise person-centred care within existing services | 2.1 – Incentivise identification of, and response to, patients’ self-identified support needs by HCPs<br>2.2 – Incentivise identification of unpaid/family carers by HCPs<br>2.3 – Incentivise identification and response to unpaid/family carers’ self-identified support needs by HCPs |
| 3. Enable identification and response to patient support needs (through evidence-based tools and approaches) | 3.1 – Support research to develop and pilot the Support Needs Approach for Patients (SNAP [37-40])<br>3.2 – Address inequities in the provision of existing referable services<br>3.3 – Increase generalists’ knowledge of non-pharmacological interventions for intractable breathlessness<br>3.4 – Prompt HCPs to establish the presence of, and review appropriateness of, patient-identified key HCPs |
| 4. Identify and support patients' unpaid/family carers (through evidence-based tools and approaches) | 4.1 – Encourage HCPs to identify and acknowledge unpaid/family carers<br>4.2 – Encourage HCPs to implement the Carer Support Needs Assessment Tool (CSNAT [32,41,42]) Approach with carers<br>4.3 – Prompt HCPs to establish the presence of, and review the appropriateness of, carer-identified key HCPs<br>4.4 – Support research to explore the feasibility of a dedicated Carer Support Nurse role [43,44] |
| 5. Identify and respond to psychological morbidity in patients and unpaid/family carers | 5.1 – Review the NICE-approved four-level model of psychological assessment and support for cancer patients and families [45] for potential adoption by HCPs working with those living with advanced COPD<br>5.2 – Provide brief, accessible, post-registration training in psychological support for practice nurses<br>5.3 – Action recommendation 3<br>5.4 – Action recommendation 4 |
| 6. Change societal attitudes and understandings of COPD, breathlessness, palliative care and unpaid/family carer support | 6.1 – Develop and test educational resources on COPD, misconceptions about people who are breathless in public, palliative care, and the caring role<br>6.2 – Encourage the provision of educational media representations of COPD, people who are breathless in public, palliative care, and the caring role |

Each recommendation is presented below in two parts: first, a narrative on the key supporting LwB evidence for the problem area, interwoven with the debate that emerged through the three linked recommendation-generating strategies and, second, a narrative on the resulting recommendation. LwB programme source studies are indicated throughout in square brackets: Longitudinal Interview Study [LIS], Care Preference Survey [CPS], or Barriers and Facilitators Study [BFS]; data collection time points are indicated for the Longitudinal Interview Study where relevant [t1-t7].

### Problem Area 1: Perseveration on the challenge of prognostication and unpredictable trajectories

HCPs found prognostication challenging and trajectories largely unpredictable in advanced COPD: they perseverated on these difficulties as barriers to meeting patient needs [BFS]. Although prognosis and trajectories are helpful drivers for appropriate care and support in cancer (where they are largely predictable) and play a role in some treatment decisions in advanced COPD, they are distracting and potentially obstructive in non-malignant long-term conditions.

Most HCPs said they would not be surprised if the patient who nominated them (through the LwB programme as their key HCP for their lung problem) died in the next 6-12 months (the “surprise question”) but added that, equally, they would not be surprised if the patient *didn’t* die in the next 6-12 months [BFS]: this is the paradox in advanced COPD. As a common entry criterion onto primary care palliative care registers the “surprise question” works well in cancer: very few LwB patients were on such registers or were discussed at related practice meetings [BFS]. Furthermore, only one of the 235 LIS patients reported receiving specialist palliative care [LIS].

HCPs felt challenged on when to start future care discussions, characterising them as needing to occur toward the end of the patient’s disease trajectory (as they did in cancer), despite acknowledging the prognostication challenge noted above [BFS]. Patient and carer preferences for discussing the future varied [LIS]. Some wanted to learn how COPD might progress, and carers particularly wanted to know what support might be available (as opposed to discussing “end-of-life” or “Advance Care Planning”). Others (usually patients) were reluctant to have such conversations, preferring to live day-to-day [LIS]. The most common unmet support need identified was with “knowing what to expect in the future”: this was identified by over a third of patients (36%, n=52/146 [LIS-t5: as t5 was when this question was first asked]) and more than half of carers (carers 53%, n=60/113 [LIS-t1]).

Finally, there is a tendency in policy documents and directives to equate palliative care with specialist palliative care, and palliative care with end-of-life care, through using these terms interchangeably. This conceptual confusion is unhelpful.

### To address Problem Area 1, the resulting Recommendation 1 is to stop focusing on the challenge of prognosis and unpredictability of trajectories as barriers to meeting needs

#### Recommendation 1 narrative

HCPs should be enabled to think differently in advanced COPD, to think beyond the “surprise question”. Prognostication-based criteria for access to appropriate care and support (rather than for making treatment decisions) addresses the needs of systems, institutions and professionals, not the needs of patients or their unpaid/family carers. Someone with advanced COPD might benefit from appropriate support or intervention regardless of whether they can be located at a point on a trajectory, and regardless of whether their need is COPD-related. Drivers for access to appropriate care and support should be person-centred, rather than prognostication- or disease-centred: perceived need is individual, not trajectory- or disease-specific.

The classic trajectory for organ failure [12,27] implies a recognisable, predictable decline in functioning, with dips from which patients only partially recover, suggesting that trigger points exist for intervention. However, such triggers may only enable reactive responses to crises rather than proactive identification and response to support needs. An alternative, proactive, person-centred approach should be adopted across health care encounters (both settings and timepoints). If care is person-centred then appropriate care and support, including access to (specialist) palliative care, should follow as a response to need rather than as a response to a trigger or a point on a trajectory.

To address the difficulty of not all patients wanting “end-of-life care discussions”, or to talk about “Advance Care Planning” (terminology derived from the cancer model), “thinking about the future” and “planning ahead” may be more engaging terms for patients and carers living with non-malignant disease and may feel less challenging for HCPs. Changing our language may also enable earlier conversations, before disease becomes too advanced. Similarly, “symptom control”, “support needs” or “supportive care” may be more engaging (and understandable) terms for patients with non-malignant disease and their carers than “palliation”. Using non-specialist terms may also help generalists consider such care as within their remit and facilitate integration of palliative care with respiratory management across professions. Table 2 outlines Action Points to enable delivery of Recommendation 1.

### Problem Area 2: Care is predominantly reactive, target-driven, led by organisational and medical agendas, with limited evidence of person-centred care

The organisation of health care rarely enables identification and response to unmet support needs, and health care systems do not routinely incentivise it. In particular, primary care annual reviews for COPD patients driven by financial incentives of the Quality and Outcomes Framework (QOF), do not incentivise person-centred care in COPD. In describing annual reviews, primary care HCPs characterised them as prioritising spirometry, medication review, and assessing breathlessness (MMRC dyspnoea scale) and oxygen saturation [BFS]: these were all QOF targets at the time of data collection along with maintaining a register of COPD patients and providing influenza immunisation [46]. Some practices had developed electronic templates to record these tasks [BFS], but the role of some of this activity for primary care management of advanced COPD was unclear e.g., primary care use of spirometry results, noting that spirometry can be uncomfortable for patients and is time-consuming for HCPs [LIS/BFS].

Thus, annual reviews, which have the potential to be proactive, appeared target-driven and task-oriented, and perceived by some patients as having little relevance to them (although some found them reassuring) [LIS]. Primary care HCPs identified non-attendance at reviews as a barrier to meeting patients’ needs, but one HCP felt that non-attendance meant the patient was probably managing well [BFS]. Patient-perceived irrelevance was not identified by HCPs as a reason for non-attendance [BFS]. These reviews are a missed opportunity to proactively identify, and respond to, patient and carer support needs, and explore preferences for future care.

Other primary and secondary care contacts were valued by patients but characterised as largely reactive and brief. Reactivity in acute exacerbations is entirely appropriate but, again, we found little reference in HCP interviews to identifying or responding to patient support needs, very little knowledge of who patients’ carers were, and even less knowledge of carers’ support needs [BFS].

Where carers were spoken of this was usually in terms of how they could support the patient [BFS].

Patients’ experiences of hospital admissions varied: some reported problems with discharge planning e.g., promised district nurse visits [LIS]. Access to secondary care outpatient appointments varied, as did patients’ and carers’ perceptions of their value [LIS]. Some liked the reassurance these contacts provided [LIS], but others questioned their purpose:

> *“…every time I go and see [consultant] it’s the same performance: blowing into that thing […], answering a few questions, then I go on my merry way. And then I go back again […] go through the exact same procedure again […]. When the [Community Respiratory Nurse] comes to see me […], same situation: check me out, blood pressure […], give me a tube to spit into […] and that is the extent of what I get. […] I haven’t seen anybody that I could actually go away from and say that they have helped me […] I know that I’m not going to get anything that’s going to relieve my problems […] I’ve never had anything like that, ever. […] It’s routine […] it’s the same old thing […]. Nothing is any different – even to the little jokes we have together […] I’ve got it off by heart”* [Patient 004-675 LIS t1]

Some respiratory consultants spoke of the importance of talking and listening:

> *“when we meet patients for the first time, we’re very good at listening […] to try and unpick what’s going on […] letting patients talk and listen to the story of what’s happened”* [Respiratory Consultant 107-006 BFS]
>
> *“sometimes 10 minutes of time of listening and saying ‘well, why don’t you try this or tweaking this?’ […] is just as valid as ‘here we go, here’s an inhaler’, but I don’t think commissioners and managers see it that way’’* [Respiratory Consultant 140-006 BFS]

Appointments, however, remained characterised as organisational and medical, even by consultants themselves:

> *“as doctors we have a […] scientific approach to things: we need to work out why you’re breathless and tell you why you’re breathless, but that doesn’t really matter to him why he’s breathless, it’s almost irrelevant”* [Respiratory Consultant 132-006 BFS]
>
> *“So there’s this problem, we can address it with this drug, those sorts of things: it’s almost a tick, tick, tick, yeah, OK’’* [Respiratory Consultant 101-400 BFS]

What patients and carers wanted were opportunities to talk to HCPs who knew them, had insight into their condition and time to talk [LIS]. Mostly they wanted reassurance and to better understand what might happen: how much worse their condition might get, what resources they might access and how. They wanted information to help them live as well as they could [LIS]. Some patients commented on the therapeutic effect of our research interviews: as researchers we were interested in their condition, had insight into what it was like to live with, we actively listened and were able to give them time to talk:

> *“I go to see a health care professional […] only when I have a lung function test, and they say ‘breathe into this’ [puffs] and they go ‘OK, you’re fine, goodbye’. And that’s it. […] I’ve never been to see anyone where they say ‘let’s talk’, you know, like I talk to you. […]. With [the GP]*
>
> *I only go to see him if I’m in a desperate situation. If I’m not in a desperate situation, I don’t waste his time […] I never talk to anybody. Nobody’s said ‘right, let’s sit down and have a chat about it’ I’ve had no support. Except for you – talking to you about it”* [Patient 022-500 LIS t5]

HCPs across settings and professions also commented on how LwB study participation had enabled reflection on their practice in relation to meeting support needs:

> *“You’re making me wonder, actually, whether I’ve got [community services] involved with him before? Whether he’d benefit? I’ll think about that the next time I see him”* [Respiratory Consultant 101-400 BFS]

### To address Problem Area 2, the resulting Recommendation 2 is to change targets to incentivise person-centred care within existing services

#### Recommendation 2 narrative

Care is often target-driven, led by organisational and medical agendas, with limited evidence of person-centred care. We should revise targets, such as QOF, for advanced disease to enable person-centred care by incentivising the identification and response to patient and carer support needs, within existing services. Table 2 outlines Action Points to enable delivery of Recommendation 2.

### Problem Area 3: Unmet patient support needs – patients had difficulty articulating their support needs and access to support varied

When asked about the quality of care, many patients spoke positively (particularly related to times of crisis) but some responded with *“what care?”*. Despite high use, the “care” element of service contacts was not always visible to them [LIS]. As noted, we found little evidence of HCP-reported proactive assessment of patient-identified support needs in primary or secondary care, except by Community Matrons, who described having more time:

> *“I can be there for an hour going through everything, whereas I think the GPs they feel pressurised”* [Community Matron 104-401 BFS].

However, time was not the only barrier: care across settings was predominantly disease-focused rather than person-centred [LIS/BFS]. Patients have support needs related to COPD but also to co-morbidities which may affect their ability to self-manage their COPD: 97% (n=228/235) LIS patients reported comorbidities, reporting a median of four other conditions (IQR 2-5).

Importantly we found wide variation in patients’ ability to spontaneously articulate their support needs. When we asked an open question about what was the most helpful, or important, thing someone could do for them today, some patients couldn’t answer and 30% (n=60) didn’t think there was anything anyone could do or didn’t know what would help [LIS]. This was despite a wealth of indirect expressions of need within the LIS interviews. This difficulty in expressing need has huge implications for support need identification (and therefore action) by HCPs. HCPs cannot rely on patients spontaneously reporting support needs or reporting needs even in response to open questions such as “how are you?”, unless in crisis [47]. Patients need help to express their support needs.

We only systematically identified unmet support needs across the LIS cohort by introducing a set of prompts at follow-up interviews (t5+) i.e., a list of potential areas of support need with a stem question “Do you need more support with…?”. This format and list of questions was adapted (with permission) for patients for the LwB programme from the Carer Support Needs Assessment Tool [32]. Sixty-five per cent (95/147) of LIS-t5 patients then identified at least one area of unmet support need. They wanted more support with: knowing what to expect in the future (36%; 53/147), practical help (35%; 52/147), understanding their illness (35%; 52/148), looking after other health problems (35%; 52/148), managing symptoms and medications (30%; 44/147), knowing who to contact if they were concerned (29%; 43/147), dealing with feelings and worries (27%; 40/147), and equipment to help them (27%; 40/147) [LIS-t5]. These unmet support needs existed despite (or because of) living with COPD for a median of 10 years [LIS]. When we asked which of the self-identified need was their greatest priority most said “understanding their illness” and “knowing what to expect in the future” [LIS] – that “future care” topic noted in Problem Area 1 that HCPs found so difficult to know when to raise with patients [BFS].

Patients also varied in their ability to answer questions about their need for named services, often asking what the named services were for: their knowledge was limited [LIS]. Frustrated by the time-limited nature of services they knew and valued, such as pulmonary rehabilitation, patients wanted maintenance courses, particularly for exercise [LIS]. Similarly, HCPs across professional groups and settings identified the limited availability of referable services as problematic e.g., pulmonary rehabilitation, clinical psychology, and community specialist respiratory nurses [BFS]. This was commented on by one respiratory consultant in his surprise at being nominated for the study by his patient as their key HCP:

> *“what it reflects is the disappointing lack of community respiratory services […] we have much less now than we had ten years ago through, I think, a disappointing view of these matters from commissioners in the area. So, I shouldn’t be his ‘key healthcare professional’ […] but, nevertheless, […] can understand that he may have a confidence in someone who’s seen him on a continuing basis over the years’’* [Respiratory Consultant 101-503 BFS].

When asked to identify which HCP gave them the most help and support with their lung problem patients predominantly identified community-based HCPs, usually their GP (34%; 78/230), community respiratory nurses (18%; 42/230), nurse practitioners or practice nurses (10%; 23/230) [LIS]. Twelve per cent (27/230) identified a hospital doctor. However, nearly a fifth (19%; 44/230) were unable to identify a “key” HCP for their lung problem, despite its longevity – not even their GP [LIS].

Breathlessness was universally experienced [LIS], but just 21% had a handheld fan – an evidence-based intervention for intractable breathlessness [48,49]. Only one patient (of 235 responding) had been referred to a specialist breathlessness intervention service, and only one to specialist palliative care [LIS]. Yet patients reported being told “there is nothing more I can do for you” by GPs and respiratory consultants, and patients’ interpretations of this varied [LIS].

Advanced COPD patients are not homogeneous and so may not fit pathways: in the event of an exacerbation, their preferences for care were influenced by the severity of their condition and support available from unpaid/family carers [CPS]. We identified wide variation in functional trajectories using validated tools [29–31], but most described their lung problem as deteriorating and feeling more breathless 18 months from baseline [t7], and worried about disease progression [LIS]. Exceptions were patients who had lost weight, stopped smoking, or recovered from an exacerbation: they reported improved symptom control [LIS].

### To address Problem Area 3, the resulting Recommendation 3 is to enable identification and response to patient support needs (through evidence-based tools and approaches)

#### Recommendation 3 narrative

Using an evidence-based designed-for-purpose prompt list in clinical practice (as opposed to often used patient reported outcome measures identifying disease impact rather than need) could facilitate patients’ expression of support needs. This could enable person-centred needs-driven conversations with HCPs to address identified needs together. The Support Needs Approach for Patients (SNAP: outlined in Box 4) [37–40], which uses this approach could be adopted (as a minimum) in community generalist settings where most service contacts in advanced COPD occur [LIS].

###### Box 4: Support Needs Approach for Patients (SNAP)

The Support Needs Approach for Patients (SNAP [37–40]) is modelled on the Carer Support Needs Assessment Tool Intervention (CSNAT-I [41]): an approach to clinical practice-based needs assessment developed to identify and respond to carers’ needs [50].

SNAP is a person-centred process of assessment and support providing patients with an opportunity to consider, express, and prioritise their support needs. It is a process facilitated by HCPs but led by the patient. Support needs are identified by the patient using a brief structured 15-item tool (the SNAP Tool). The patient-completed SNAP Tool then enables a conversation between the patient and their HCP to prioritise and respond to the patient-identified needs together. The SNAP Tool consists of 15 broad validated evidence-based support domains and asks the patient whether they need more support with each e.g., “understanding your illness” or “aids or equipment to help you”. It therefore directly identifies patient support needs.

The SNAP Tool development was grounded in data from patients with COPD [37], validated in patients with COPD [38] and has been piloted in patients with COPD [39]. The SNAP Tool is brief and simple, yet comprehensive. Existing needs assessment tools are too long and predominantly developed for cancer or for screening for referral to specialist palliative care. SNAP could enable HCPs to deliver on the policy rhetoric of person-centred care by facilitating identification of support needs, letting patients know it is appropriate to ask HCPs about their concerns (by acting as a prompt, implicitly giving patients “permission” to raise issues), and making care more proactive (by starting conversations between patients and HCPs on patient-identified and prioritised areas of support need).

SNAP and CSNAT-I are a change from usual practice where identifying need is largely HCP-led thus, like CSNAT-I, introducing SNAP to HCPs requires training [51].

SNAP was developed in direct response to Recommendation 3 of the LwB programme and was subsequently included in the British Lung Foundation led Taskforce on Lung Health [21] and the Royal College of General Practitioners and Marie Curie UK’s General Practice Core Standards for Advanced Serious Illness and End of Life Care, known as the Daffodil Standards [52].

It has a dedicated website that includes online training and licensing access [40]

Responses to identified support needs will vary, but conversations about unmet support needs may be an intervention in itself. Needs-driven talk may make care and support more visible, enhancing clinician-patient relationships: “talk” was something these patients wanted [LIS]. Responses may also include signposting or referral on but, as noted, we need greater equity in provision of highly valued referable services. We need also to increase HCP awareness of local specialist services e.g., only one LIS cohort patient had access to the Cambridge Breathlessness Intervention Service [53].

Improving generalists’ knowledge, and delivery of, evidence-based non-pharmacological interventions for intractable breathlessness could extend responses to patient-identified need e.g., the handheld fan [48,49]. This may also reduce HCPs’ feelings of helplessness in managing advanced COPD, reducing those noted “there is nothing more I can do for you” messages.

It may be useful for HCPs to establish if patients can identify the key HCP who supports them (and advise on/review that choice) and liaising with them as required. Finally, identifying and addressing support needs should be ongoing as patients’ circumstances change: our patients became widowed, moved house and either gained a carer for the first time, lost their carer, or experienced carer change during their 18-month follow up [LIS/CPS]. Table 2 outlines Action Points to enable delivery of Recommendation 3.

### Problem Area 4: Unmet carer support needs – little evidence of identification of unpaid/family carers, or their support needs

Most patients reported having an unpaid/family carer (88%, n=205/235 [LIS]). Some had more than one and for some this person changed during the 18-month follow-up. A quarter were supported by their carer for over 50 hours a week (25%, n=48/189) and just over a quarter lived separately from them (28%, n=57/205). Patients reported that most carers were spousal (63%, n=125/200), giving daily help (74%, n=144/195) with practical tasks such as shopping (62%, n=139/225), housework (52%, n=118/228), cooking (43%, n=98/228), transport (43%, n=97/226), and paperwork and phone calls (28%, n=63/227), and personal tasks such as walking (28%, n=62/226), washing (26%, n=58/226), dressing (22%, n=49/226), and helping during the night (13%, n=29/224) [LIS].

A third of carers reported having other caring responsibilities (34%, n=38/113), usually for other adults [LIS]. Carers were not just caring for patients in relation to their COPD: carers identified other health problems their patients had which impacted on, or required, their caring role [LIS]. Carers had their own physical health problems, some of which impacted on their ability to support the patient [LIS] including back pain (18%, n=19/104), arthritis (12%, n=12/104) and mental health problems (8%, 8/104).

Carers felt unprepared for many aspects of their role (LIS: PCS [33]) and had a range of unmet carer support needs (LIS: CSNAT [32]). The median duration of their caring role was seven years (IQR: 3-12 years), usually increasing in intensity over that time. These carers felt unprepared, and had unmet support needs, despite (or sometimes because of) the longevity of their role [LIS].

Two thirds of carers could not identify a HCP who supported them in their role (65%, n=67/103). Those that could predominantly identified a community-based HCP: usually their GP (n=21/103), a community respiratory nurse (n=7/103), Community Matron (n=5/103), nurse practitioner or practice nurse (n=2/103); one unpaid/family carer named a hospital consultant [LIS]. There was an unchecked assumption among HCPs that carer support was covered by others [BFS]: *“Clearly it’s important, but… I would naturally assume that those things would be being dealt with on a community basis’’* [Respiratory Consultant 101-400 BFS].

HCPs rarely knew who the carers were, and their needs were rarely identified or responded to [BFS]. This held true across professional groups and settings (primary and secondary care), with the exception of Community Matrons. One respiratory nurse said: *“I never […] have that contact with her and [the patient] just mentions her name, I don’t know what her support needs are”* [Nurse Practitioner 107-101 BFS]. One GP described his role with carers as *“an indirect one”* [GP 102-40 BFS]; another noted that *“in the thick of a busy consultation [carers] can sometimes be easily forgotten”* [GP 122-400 BFS].

Respiratory consultants who reported engaging with carers did so in order to determine patient need, rather than the carer’s own support needs:

> *“part of the consultation is always to listen to him and then to introduce why he might not be OK, by listening to his wife. […] she will pipe up and say ‘this is what’s happened’. Now sometimes patients’ carers will over-report and you wonder where you’re at, but I think she’s reliable – it would be a worse consultation if she wasn’t there”* [Respiratory Consultant 117-006 BFS]

### To address Problem Area 4, the resulting Recommendation 4 is to identify and support patients’ unpaid/family carers (through evidence-based tools and approaches)

#### Recommendation 4 narrative

Unpaid/family carers provided considerable practical, personal and emotional support to patients but had their own unmet support needs: both to help them provide care and for their own physical/mental health and wellbeing [LIS]. Carers are patients too. We need to be considerably better at identifying, acknowledging and supporting them. We need to start by identifying who patients’ carers are – this is the first step in proving support [54]. However, cognisant that not all patients (or carers) understand or accept the label “carer”, HCPs should ask patients: *“who gives you help and support at home?*” rather than *“do you have a/who is your carer?”* [54].

HCPs should establish if carers have a key HCP who supports them in their role (and advise on/review that choice) and liaise with them as needed. We should also acknowledge and support that some patients are also carers themselves, providing care and support to other family members and friends.

We should treat patient-carer dyads as a unit of care in advanced disease. Carers could be invited to patients’ annual reviews in primary care; some pulmonary rehabilitation programmes already encourage carer attendance. Within such settings, using the CSNAT Intervention [32,41] (which has been updated and validated for COPD carers [42]) could facilitate identification and response to carer needs.

To address the lack of preparedness to care, we should develop and deliver evidence-based carer learning resources relating to both carers’ own needs [55] and those of the patient. And to address the limited evidence of carers active help-seeking [LIS], we should consider the utility of a dedicated Carer Support Nurse role to support more vulnerable carers and raise the profile of carers among other HCPs [43,44]. Table 2 outlines Action Points to enable delivery of Recommendation 4.

### Problem Area 5: Patient and carer psychological morbidity

Around half of the patients self-reported anxiety or depression (55%, n=124/227 and 48%, n=110/230 respectively [LIS]), but 37%-43% of these had not told a HCP about this (37%, n=43/115 and 42%, n=44/105 respectively [LIS]. They felt there was little HCPs could do and they *“just had to get on with it”*. However self-reported anxiety and depression were positively associated with Hospital Anxiety and Depression Scale scores (HADS) [30] [LIS]. HADS scores were also higher than population norms for both patients and carers (Box 3 [LIS]) [56]. Further, the study’s Depression Protocol was actioned for six patients and two carers participating in the LIS component i.e., GPs or Community Matrons were notified of over-threshold HADS-D scores, or of researcher concerns, with the permission of the patient/carer.

Patients powerfully described feelings of frustration, anger, worry, panic, guilt, failure, being a burden, identity loss, lack of knowledge, impacts of limitations, reliance on others, boredom, and the psychological impacts of house-boundness [LIS]. Just 5% of patients (n=11/227 [LIS]) had accessed peer support groups: availability of these groups varied. Some carers described the particular challenge of managing patient anger and frustration [LIS].

Some HCPs (particularly nurses and hospital consultants) identified limitations in their own skills in directly meeting the psychological needs patients [BFS]. Nurses lacked confidence in psychological support, whereas hospital consultants spoke of referring on:

> *“If it’s an area that is really not my expertise, or I feel the GP would have better expertise, let’s say […] they return to clinic, I believe that their chest is stable, but they appear down, demotivated, and I think that they’ve changed, then rather than tackling that myself, I will usually flag it to the GP, that I think there’s been a new change, could they please assess and see what they think”* [Respiratory Consultant 117-006 BFS]

### To address Problem Area 5, the resulting Recommendation 5 is to identify and respond to psychological morbidity in patients and unpaid/family carers

#### Recommendation 5 narrative

Routinely asking patients and their carers if they are bothered by anxiety or depression could identify those with potential psychological morbidity who might benefit from support. Structured screening tools, such as the HADS [30], are often used in clinical practice but should be scored and responded to where patients or carers score over thresholds.

Psychological assessment and support are not just the remit of specialists. We should enable generalists to assess psychological health, provide non-specialist psychological support where appropriate, and recognise and refer-on when specialist intervention is required. Reviewing, adapting and adopting recommendations such as the NICE-approved four-level model of psychological assessment and support for cancer patients and their families [45] may provide structured guidance for generalists in COPD. In order to deliver Level 1 psychological support (recognition of psychological needs followed by effective information giving, compassionate communication and general psychological support) [45], we should routinely provide brief, accessible, post-registration training in psychological support for generalists such as practice nurses e.g., Sage & Thyme [57]. Table 2 outlines Action Points to enable delivery of Recommendation 5.

### Problem Area 6: Perceptions and attitudes

When telling their story most patients described COPD as something they had brought on themselves through smoking; many expressed guilt about family impact and their need for scarce NHS resources [LIS]. A minority reported occupational or genetic causes for their COPD but suffered the common assumption (and judgement) that their condition was self-inflicted [LIS]. Patients reported embarrassment when breathless or coughing in public, describing the impact of negative reactions of others [LIS].

Patients commented on the high profile of cancer compared to COPD both for raising awareness (to enable prevention) and for raising money for research and supportive services [LIS]. Patient and carer understanding of the progressive nature of COPD was sometimes limited, and some reported limited knowledge among wider family and friends [LIS]. Many equated palliative care and hospice care with end-of-life care, presuming that referral to such services meant they were actively dying [LIS].

Carers were often surprised when we asked them which HCP gave them the most support with their caring role [LIS]: they have been referred to elsewhere as “hidden patients” [58] reluctant to ask for help, placing their patient’s needs as paramount.

### To address Problem Area 6, the resulting Recommendation 6 is to change societal attitudes and understandings of COPD, breathlessness, palliative care and unpaid/family carer support

#### Recommendation 6 narrative

Given the pervasive effects of guilt and stigma on patients and their families, we should facilitate change in societal attitudes to COPD and other smoking-related conditions, and responses to people who are breathless in public. We also need better societal understanding of the progressive life-limiting nature of COPD. This could facilitate conversations about future care between both patients and HCPs, and patients and carers.

We need to improve societal understanding of the timing, goals and role of palliative care in general. As noted under Recommendation 1, policy documents and directives often (unhelpfully) use terms interchangeably, equating palliative care with specialist palliative care, and palliative care with end-of-life care.

And as a society, we need to understand the fragility of the caring role, acknowledging that unpaid/family carers sometimes want to stop caring or want someone else to take charge. We need to ensure they can ask for help.

These ambitious goals could be addressed through public education about COPD, about misconceptions about people who are breathless in public, about palliative care, and about the caring role. This might be achieved through media representations such as those sometimes seen for cancer (e.g., soap opera and medical drama storylines), documentaries, and advertisements for relevant charities and peer support groups across a range of media. Table 2 outlines Action Points to enable delivery of Recommendation 6.

### Stakeholder survey responses to the recommendations

Responses were received from 83 stakeholders, including allied health professionals, doctors and nurses, working across primary and secondary care [36]. Overall, there was high agreement that all six recommendations would improve care and support for people with advanced COPD, however, there was caution around the ease of implementing some, which varied by professional group and setting. Concerns largely related to the time and resources for implementation and resources needed to respond appropriately to any unmet needs identified [36].

## DISCUSSION

As the largest population-based, mixed method, multiple-perspective, longitudinal study of care and support in advanced COPD conducted in the UK at the time, the Living with Breathlessness Study programme sought to inform a new paradigm to improve care and support in advanced COPD. At the outset we envisaged recommending large-scale initiatives such as the formulation of pathways, or reconfiguring existing services into integrated packages of care, or developing new services. Had we collected single perspective, single method, cross-sectional data our findings may have led to such recommendations. However, the uniqueness of this study lay in its breadth and depth: its multiple-perspective, mixed method, longitudinal approach uncovered a range of experiences (both between individual patients and between localities) and divergence in expectations and experiences between the multiple perspectives explored.

In brief, the study identified that HCPs perseverated on the challenge of prognosis and unpredictability of trajectories as barriers to meeting needs. Care was predominantly reactive, target-driven, led by organisational and medical agendas with limited evidence of person-centred care. There were unmet patient support needs which patients rarely articulated. There were unmet carer support needs and even little evidence of identification of carers. Patient and carer psychological morbidity was higher than population norms and not always reported to HCPs: patients’ self-identified that their key HCPs lacked skills and confidence in managing psychological morbidity. Patients and carers reported negative societal attitudes to COPD and breathlessness and held misconceptions of palliative care and carer support.

National stakeholders advised that our findings confirmed some of what was known (reflecting both their practice experience and earlier literature), but importantly highlighted areas where there has been a failure to act on previous findings: our six evidence-based recommendations provide a route map for addressing these issues. For example, our finding that primary care annual reviews appeared largely target-driven and task-oriented is supported by others [18,19], but our actions points propose novel mechanisms to respond to this. A limitation of the study is that the data were collected, and stakeholder consultations conducted (workshop and survey) 10 years ago, between 2013 and 2016.

The six resulting recommendations remain relevant in seeking to enable delivery of person-centred care by facilitating the required paradigm shift within HCPs and multiple levels of systems i.e., through person-, system-, and societal-level initiatives. This paradigm shift is from the one that is driven by the cancer model, that is prognosis and trajectory-oriented, and that is led by disease-specific organisational and medical agendas, to one of person-centred care. Person-centred care is not a new concept [59] but the six recommendations, grounded in patients’ and carers’ experiences, may be novel in explicitly identifying evidence-based ways to help patients, carers and HCPs achieve the rhetoric of person-centred care within existing services and platforms, addressing the lack of guidance on how person-centred care can be achieved. This includes adopting evidence-based approaches and tools across settings, services and localities to help HCPs, patients and carers start conversations to enable better care and support. Three evidence-based interventions arose directly from the LwB programme: the Support Needs Approach for Patients (SNAP [37–40]), the updating of CSNAT to ensure relevance for COPD carers [42], and the Carer Support Nurse role [44].

Enabling person-centred care through these mechanisms may make patients and unpaid/family carers feel better supported, may increase the visibility of care and support, may avert crises, and may make HCPs feel less helpless – which together could lead to both financial and impact savings.

The study’s findings are important for those living with advanced COPD but stakeholders advised that the resulting six recommendations have resonance for patients and unpaid/family carers living with COPD which is not yet advanced, and generic resonance for other similar advanced non-malignant indeterminate long-term conditions which are often accompanied by a high number of co-morbidities: for patients and unpaid/family carers who are living in between conventional disease management and end-of-life care, and for those HCPs striving to support them.

## CONCLUSION

In conclusion, the comprehensive Living with Breathlessness Study programme provided an exceptional view of the experience of people with advanced COPD, and those supporting them (unpaid/family carers and HCPs). We propose that enactment of the resulting six evidence-based recommendations through their action points could inform a new paradigm to improve care and support in advanced COPD and hold potential for other patients and unpaid/family carers who are living in between conventional disease management and end-of-life care, and for those HCPs striving to support them.

## ACKNOWLEDGEMENTS

The authors particularly thank: all participating patients, unpaid/family carers, HCPs and stakeholders; the former Primary Care Research Networks and all recruiting practices (East of England and South London); British Lung Foundation Breathe Easy groups and additional patient and carer representatives for their PPI roles; Dr Silvia Mendonca for statistical advice; Dr Stephen Barclay for enabling BFS component delivery; Lauren Milden for dissemination event planning; Kevin Houghton and Sam Barclay for administrative support and data entry; and Dr Sara Booth, Dr Patrick White, Sophie Howson, Dr Ravi Mahadeva, Professor Gunn Grande and Professor Chris Todd as Living with Breathlessness Programme Advisory Group members.

This paper is dedicated to our colleague and friend Dr Sara Booth, who died on 10 September 2022, leaving an extraordinary legacy: her BMJ Obituary describes her life and work [60]. As an international expert on breathlessness, Sara made a considerable contribution throughout the study as a Living with Breathlessness Programme Advisory Group member.

## FUNDING ACKNOWLEDGEMENTS

The Living with Breathlessness Study is independent research supported by a Marie Curie research grant (C28845/A14129) and the National Institute for Health Research: Professor Morag Farquhar was funded by a National Institute for Health Research (Career Development Fellowship, CDF-2012-05-218). The views expressed in this publication are those of the author(s) and not necessarily those of the NHS, the National Institute for Health Research or the Department of Health or other funders.

## DATA AVAILABILITY

The data that support the findings of this study are available on reasonable request from the corresponding author.

